# Isolation and contact tracing can tip the scale to containment of COVID-19 in populations with social distancing

**DOI:** 10.1101/2020.03.10.20033738

**Authors:** Mirjam E. Kretzschmar, Ganna Rozhnova, Michiel van Boven

**Author notes:** corresponding author: Mirjam E. Kretzschmar, Julius Center for Health Sciences and Primary Care, University Medical Center Utrecht, Utrecht.

## Abstract

**Background:** Novel coronavirus (SARS-CoV-2) has extended its range of transmission in all parts of the world, with substantial variation in rates of transmission and severity of associated disease. Many countries have implemented social distancing as a measure to control further spread.

**Methods:** We evaluate whether and under which conditions containment or slowing down COVID-19 epidemics are possible by isolation and contact tracing in settings with various levels of social distancing. We use a stochastic transmission model in which every person generates novel infections according to a probability distribution that is affected by the incubation period distribution (time from infection to symptoms), distribution of the latent period (time from infection to onset of infectiousness), and overall transmissibility. The model distinguishes between close contacts (e.g., within a household) and other contacts in the population. Social distancing affects the number of contacts outside but not within the household.

**Findings:** The proportion of asymptomatic or unascertained cases has a strong impact on the controllability of the disease. If the proportion of asymptomatic infections is larger than 30%, contact tracing and isolation cannot achieve containment for an R_0_ of 2.5. Achieving containment by social distancing requires a reduction of numbers of non-household contacts by around 90%. Depending on the realized level of contact reduction, tracing and isolation of only household contacts, or of household and non-household contacts are necessary to reduce the effective reproduction number to below 1. A combination of social distancing with isolation and contact tracing leads to synergistic effects that increase the prospect of containment.

**Interpretation:** Isolation and contact tracing can be an effective means to slow down epidemics, but only if the majority of cases are ascertained. In a situation with social distancing, contact tracing can act synergistically and tip the scale towards containment, and can therefore be a tool for controlling COVID-19 epidemics as part of an exit strategy from current lockdown measures.

**Funding:** This research was partly funded by ZonMw project number 91216062.

**Research in context:** *Evidence before this study:* As of 8 April 2020, the novel coronavirus (SARS-CoV-2) has spread to more than 170 countries and has caused near 90,000 deaths of COVID-19 worldwide. In the absence of effective medicines and vaccines, the preventive measures are limited to social distancing, isolation of confirmed and suspected cases, and identification and quarantining of their contacts. Evidence suggests that a substantial portion of transmission may occur before the onset of symptoms and before cases can be isolated, and that many cases remain unascertained. This has potentially important implications for the prospect of containment by combinations of these measures.

*Added value of this study:* Using a stochastic transmission model armed with current best estimates of epidemiological parameters, we evaluated under which conditions containment could be achieved with combinations of social distancing, isolation and contact tracing. We investigated the level of social distancing needed for containment, and how an additional implementation of isolation and contact tracing may likely help to in reducing the effective reproduction number to below 1, the critical threshold. We analyzed what proportion of household and non-household contacts need to be isolated effectively to achieve containment depending on the level of social distancing in the population. We estimated the impact of combinations of these measures on epidemic growth rate and doubling time for the number of infections. We find that under realistic assumptions on the level of social distancing, additional isolation and contact tracing are needed for stopping the epidemic. Whether quarantining only household contacts is sufficient, depends on levels of social distancing and timeliness of tracing and isolation.

*Implications of all the available evidence:* Our analyses based on best understanding of the epidemiology of COVID-19, highlight that if social distancing is not complete, isolation and contact tracing at least of household contacts can help to delay and lower the epidemic peak. High levels of timely contact tracing of household and non-household contacts may be sufficient to control the epidemic.

## Introduction

As of early April 2020, the number of infections of the novel coronavirus (SARS-CoV-2) is still increasing at an exponential rate in many countries, while the virus is expanding its range to all parts of the world. There are no registered effective medicines, treatment options are mainly supportive, and there are no vaccines available, limiting preventive measures mainly to social distancing combined with isolation of infected persons and those that have high likelihood of being infected, for instance because they have been traced as contacts of infected persons [1, 2]. However, recently it was questioned by high-ranked officials of the World Health Organization (WHO) whether social distancing or lockdowns are sufficient to stop the SARS-CoV-2 outbreak (https://www.reuters.com/article/us-health-coronavirus-who-ryan/lockdowns-not-enough-to-defeat-coronavirus-whos-ryan-idUSKBN2190FM). Possibly, additional measures are needed, for example active tracing of contacts in combination with isolation of infected contacts. Also, such measures will be important in the context of exit strategies, i.e. once social distancing measures are reduced or lifted, as has been suggested recently [3]. It is unclear, how effective such combinations of interventions can be in populations with social distancing in place.

To what extent local containment or local slowing down of an epidemic by isolation and contact tracing is successful depends on the fraction of infections that remain asymptomatic or have mild disease, and on the infectiousness before the onset of symptoms [4, 5]. It is known that a high probability of asymptomatic infection, a high proportion of transmission occurring before the onset of symptoms, a long delay between case finding and isolation, and high overall transmissibility all factor in negatively in the likelihood that an outbreak can be contained [6-9]. For SARS-CoV-2, evidence indicates that a high fraction of infected persons is infectious before they show symptoms (up to 50%), that a substantial fraction of infections may be asymptomatic or show only mild symptoms (up to 80%), and that the epidemic doubling time in the absence of interventions is approximately one week or perhaps even less [5, 10-16]. On the other hand, it is also reported that with intensive contact tracing it may be possible to trace the majority (>80%) of secondary infections [17].

Here we provide model-based analyses of the impact of isolation and contact tracing in a setting with various levels of social distancing measures, using varying levels of the effectiveness and timeliness of contact tracing. It is important to consider the impact of each of these interventions in isolation but also in combination, as it is well known that each intervention that reduces transmission in the population (i.e. reduces the reproduction number) is expected to increase the effectiveness of additional interventions in a synergistic manner [18]. We show how those measures impact on the cumulative numbers of infecteds during an outbreak and compare with recent outbreak data from various countries [19].

We focus on conditions that make containment of an early epidemic possible, but also on the impact of isolation and contact tracing when containment is not possible, or in an exit phase from social distancing. In a population with social distancing in place, we investigate what the additional impact of isolation and contact tracing can be for reaching containment or slowing down the spread. We report effective reproduction number, the (exponential) rate of increase, and the doubling time of the epidemic for scenarios with various combinations of interventions. Considering that the capacity of healthcare systems is limited, it is important to assess which interventions are most effective in slowing down the rate of increase of healthcare demand during an ongoing outbreak. As it is likely that, on the one hand, isolation and contact tracing will be more effective in close contact settings with well-defined contacts (household, workplace) than in the community (commuting, public spaces), while, on the other hand, the potential impact of household interventions on the epidemic could be smaller, we stratify the analyses by transmission setting (henceforth called household and non-household) [8]. As many of the relevant epidemiological and intervention parameters are still quite uncertain, or may be variable in different settings, we focus throughout on a systematic analysis of the relation between key parameters for timeliness and completeness of contact tracing and main outcomes such as the effective reproduction number and the epidemic growth rate.

## Methods

We modify a model that was developed earlier for similar aims in another context [8]. The stochastic model describes an epidemic in its early phase as a branching process. Starting from a small set of initially infected individuals, the model calculates the numbers of latently infected persons, infectious persons, and persons that are diagnosed and isolated in time steps of one day. Latent infection, infectivity during the infectious period, and daily contact rates are quantified using distributions taken from the literature (Table 1). We distinguish between household contacts and non-household contacts, which differ in the risk of infection and the delay and effectiveness of tracing and isolation. Intervention effectiveness is determined by the daily probability of being diagnosed during the infectious period (Table 2). Furthermore, intervention effectiveness depends on the delays in tracing household and non-household contacts, respectively, and the proportions of contacts can be found and isolated. We assume that isolation is perfect, i.e. that isolated persons cannot transmit any longer. The model is described by a set of difference equations, and allows for explicit computation of the basic reproduction number R_0_ and the effective reproduction number under interventions R_e_. The model was coded in Mathematica 12.1.

**Table 1:**
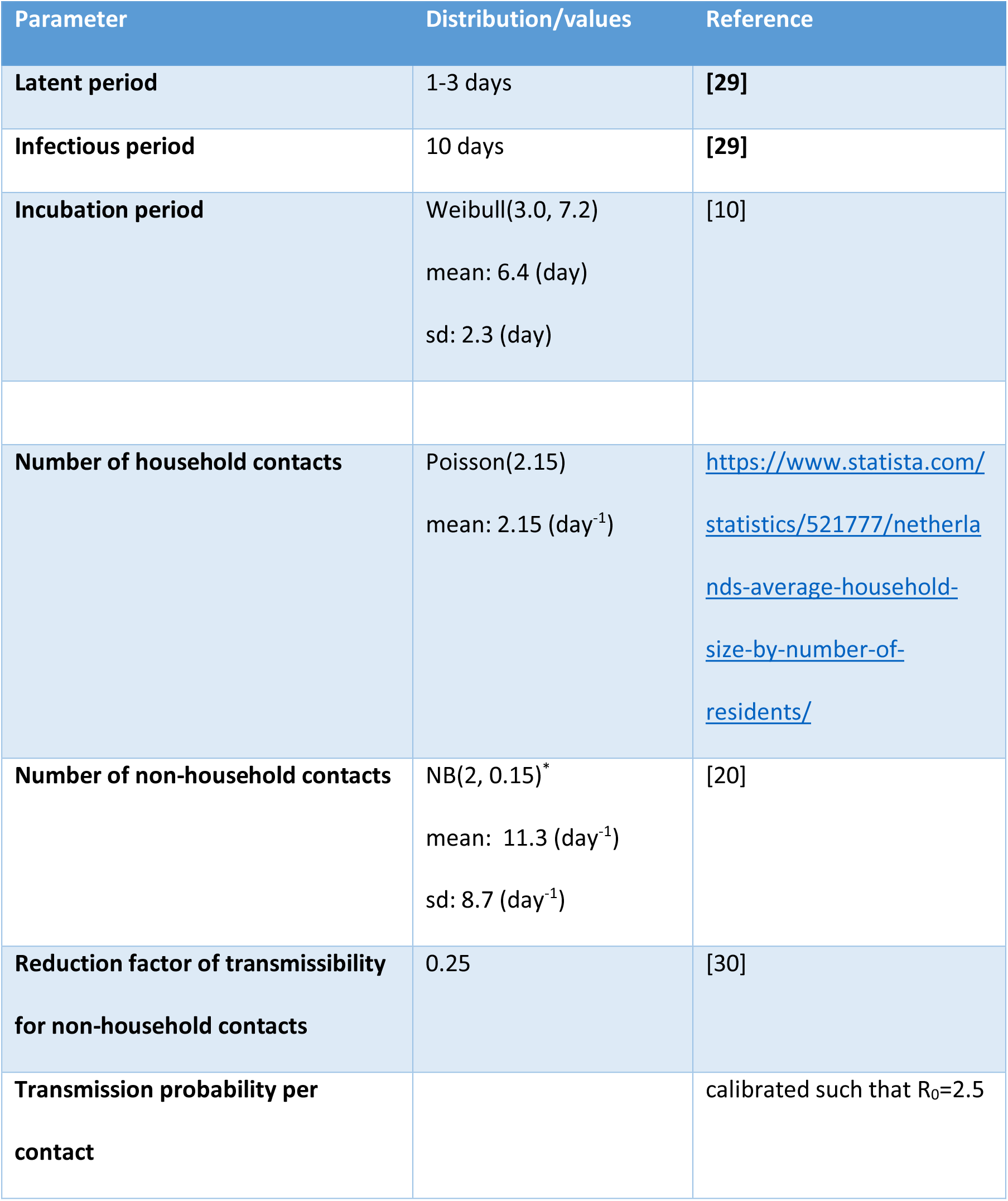
Disease and transmission related parameters.

| Parameter | Distribution/values | Reference |
| --- | --- | --- |
| **Latent period** | 1-3 days | [29] |
| **Infectious period** | 10 days | [29] |
| **Incubation period** | Weibull(3.0, 7.2)<br>mean: 6.4 (day)<br>sd: 2.3 (day) | [10] |
| **Number of household contacts** | Poisson(2.15)<br>mean: 2.15 (day<sup>-1</sup>) | https://www.statista.com/statistics/521777/netherlands-average-household-size-by-number-of-residents/ |
| **Number of non-household contacts** | NB(2, 0.15)*<br>mean: 11.3 (day<sup>-1</sup>)<br>sd: 8.7 (day<sup>-1</sup>) | [20] |
| **Reduction factor of transmissibility for non-household contacts** | 0.25 | [30] |
| **Transmission probability per contact** | | calibrated such that $R_0=2.5$ |

**Table 2:**
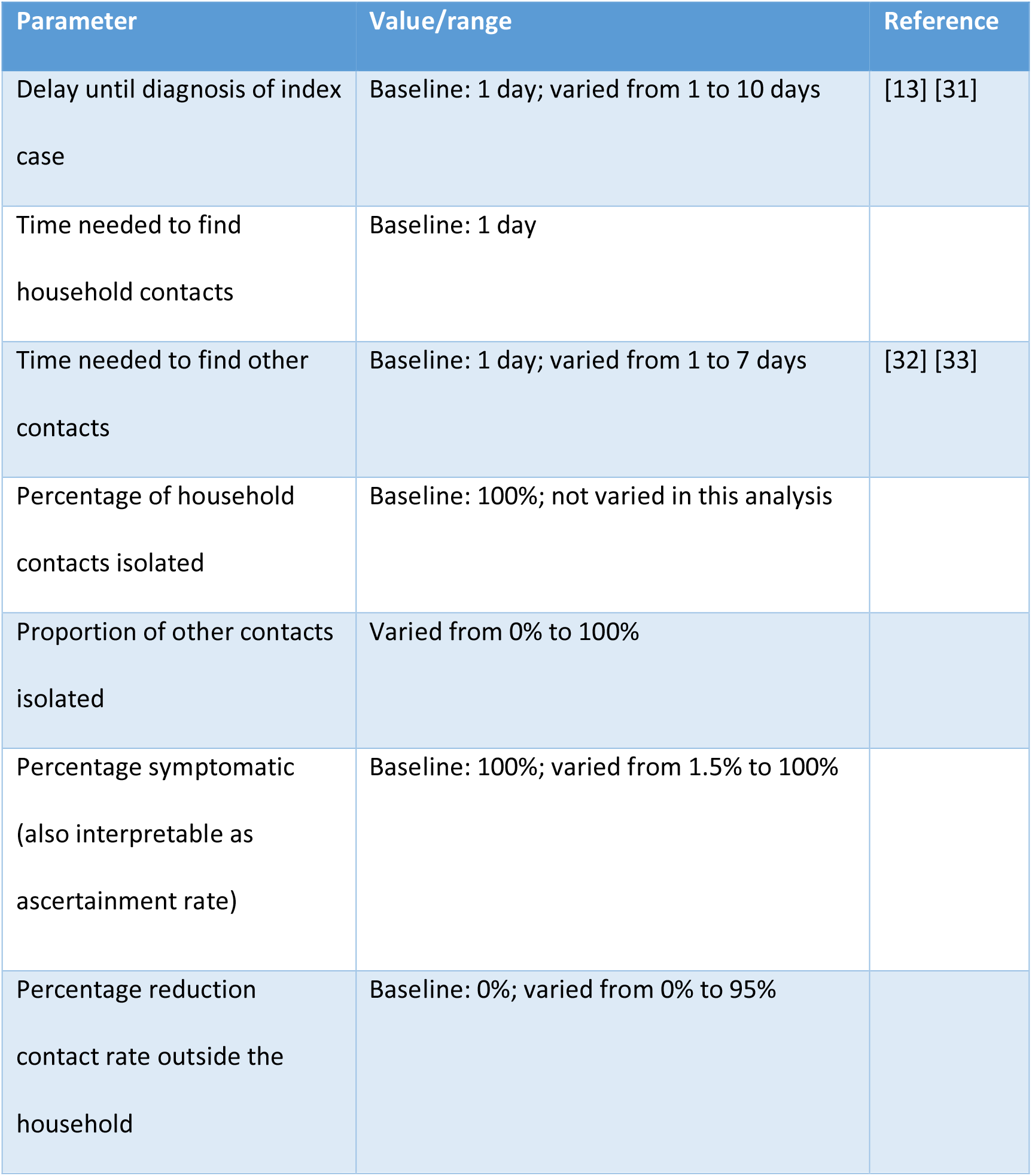
Parameters related to diagnosis and contact tracing.

### Natural history

We assume that the latent period lasts between 1 and 3 days. Individuals then become infectious for at most 10 days. Infectivity is high at the beginning of the infectious period and decays to low levels during these 10 days (Figure S1a). The probability of symptoms onset increases during the first 4 days of the infectious period, thereby influencing the daily probability of diagnosis during the infectious period (see below).

An infectious individual makes contacts with household members and persons outside the household. We model the daily number of household contacts with a Poisson distribution, and the numbers of non-household contacts with a negative binomial distribution (Table 1), with parameters based on the average household size in the Netherlands, and numbers of contacts observed in a contact study in the Netherlands (FigureS1b) [20]. With the chosen parameters, the mean number of contacts per day is 13.2 (SD 8.5).

On each day of the infectious period, an individual makes a number of contacts according to the contact distribution. This number is reduced by a factor describing the probability that the contact person has already been infected during earlier contacts with the index person. More precisely, the number of contacts is reduced by a factor *f*_*k*_ per day *k* of the infectious period, describing the probability that a contact has already been infected on previous days of the infectious period of the index case. The probability of transmission upon contact with a susceptible household contact is given by the distribution in Figure S1a. As contacts with persons outside the household are often less close, we assume that the transmission probability for these contacts is lower by factor 0.25. Figure S1c shows the percentage of onward transmissions per day of the infectious period, e.g. more than half of all onward transmissions occur during the first 3 days of the infectious period.

### Social distancing

Social distancing can be self-imposed, if people decide to reduce their social contacts during the outbreak, and it can be government-imposed by closing schools, workplaces, and other venues of social gatherings. Here we assume that when social distancing is applied, household contacts remain unchanged, but the mean number of non-household contacts is reduced. This is implemented by a reduction factor in the mean of the negative binomial distribution describing non-household contact numbers. The reduction factor for social distancing was varied between 0% and 95%. In scenarios with social distancing we assumed that only 60% of cases are symptomatic or can be ascertained.

### Diagnosis, contact tracing, and isolation

An infectious person becomes symptomatic with a given probability per day of the infectious period. For SARS-CoV-2 the probability of developing symptoms is high in the first few days of the infectious period. We assume that if an infected and infectious person has not developed symptoms by day 6, the probability that he/she will still do so is very small. The probability of developing symptoms determines whether he/she will be diagnosed and isolated. The total probability of developing symptoms determines the fraction that remains asymptomatic or otherwise undiagnosed, i.e. if the total probability of developing symptoms is smaller than 1, a proportion of the infected persons will remain undiagnosed and can transmit throughout their infectious period. With the assumed distributions, on average about half of all potential onward transmissions will have occurred before an infected person is diagnosed and isolated. The diagnosis can be delayed, which in the model is implemented by setting the diagnosis probability to zero for the number of days of delay. This delay describes the time between symptom onset until a symptomatic person visits a GP or hospital and gets diagnosed.

If an individual is diagnosed, contacts will be traced, and in case they are infected will be diagnosed and isolated. Tracing goes back in time to trace all contacts during the infectious period of the index case. There may be a delay before contacts are found and diagnosed, and only a fraction of all contacts may be found. These parameters, tracing delay and tracing coverage, may be different for household and non-household contacts. We assume that all traced infected persons are immediately isolated and cannot transmit any further.

Therefore, the only individuals who will continue transmitting are those who are not found by tracing and are not yet diagnosed.

### Baseline scenario

For assessing the effectiveness of contact tracing and isolation, we use a best case scenario, where all parameters are set to optimistic values. We assume that when a case is diagnosed, he/she will immediately be isolated and this will stop onward transmission completely. Furthermore, we assume that all contacts will be traced, and if found infected will be isolated immediately. We assume that it takes 1 day to find and isolate both household and non-household contacts. The rationale for using these overly optimistic assumptions as a baseline is that we want to investigate the maximum contribution contact tracing can provide for achieving containment. In sensitivity analyses we investigated for various control parameters at which point of diverging from the baseline parameters control of the outbreak will be lost (Supplementary Information).

### Output variables

We compute cumulative numbers of cases for a time period of 50 days after the epidemic has reached more than 100 cases. We compare doubling time of the epidemic with only isolation of diagnosed cases to situations with various levels of social distancing, isolation and contact tracing. We assume that all diagnosed cases are isolated upon diagnosis and cannot transmit further. We plot the epidemic from the time the cumulative number of infected persons is above 100. We compare these trajectories to similar trajectories for China, Germany, Italy, Japan, Turkey, and the US using data downloaded from ourworldindata.org [19]. We show what the additional contribution of isolation and contact tracing can be in the attempt to bend the curve towards longer doubling times.

The model allows an explicit calculation of the basic reproduction number R_0_ and the effective reproduction number R_e_ [8]. R_0_ is defined as the number of secondary cases an index case generates on average in a susceptible population, and R_e_ is the number of secondary infections per case when an intervention is in place. R_0_ is determined by daily transmission probabilities and numbers of contacts. The effective reproduction number is in addition determined by the level of social distancing, diagnosis probabilities, tracing delays, and tracing coverages per day of the infectious period. We can therefore investigate how R_e_ depends on R_0_ and on the intervention parameters.

We are interested in the critical tracing coverage, i.e. what proportion of non-household contacts needs to be found and isolated to control the outbreak, for populations with various levels of social distancing. Furthermore, we study the epidemic growth rate (or epidemic doubling time) without and with contact tracing and isolation and various levels of social distancing. In sensitivity analyses, we study how these quantities depend on the delay to diagnosis of cases and on the delay in contact tracing. For example, we assume that household contacts can be traced with a high coverage and without delay, but that tracing of non-household contact may take longer and be less complete.

Based on the distribution of the latent and infectious periods and infectivity, we calculate the exponential growth rate and doubling time under various assumptions on the intervention parameters. This gives additional information for situations where the outbreak is not controllable, because intervention measures will lower the growth rate and increase the epidemic doubling time.

We investigate how controllability of the outbreak depends on the fraction of infections that develop symptoms and therefore vary this percentage between 0% and 100%. We then considered combinations of interventions and their impact on effective reproduction number, growth rate, and doubling time of the epidemic. We varied levels of social distancing, and coverage of tracing of household and non-household contacts. In our analysis for different levels of social distancing we assumed that 60% of infected persons develop symptoms.

## Results

### Basic and effective reproduction numbers

In the baseline scenario without interventions we calibrate the transmission probability such that the basic reproduction number equals R_0_ = 2.5. In that case, 39% of transmissions take place among household contacts. The basic reproduction number of household contacts is 0.97, and that of non-household contacts 1.53. Hence, if all non-household transmissions could be prevented, the outbreak would be just under the control limit. In the baseline scenario without interventions the exponential growth rate is 0.19 per day and the doubling time is 3.6 days, which agrees with published estimates [13]. For sensitivity analysis of the relation between R_0_ and R_e_ for various combinations of parameters, see the Supplementary Information.

### Outbreak doubling times

Isolation of diagnosed cases alone reduces the doubling time to almost 4 days. If social distancing is implemented 28 days after the first case was diagnosed, and the number of non-household contacts is reduced by 70% [21], the doubling time will increase to around 14 days (Figure 1a). The figure also shows that it takes a week before the impact on the epidemic growth starts to be visible. If higher levels of social distancing can be achieved, the doubling times can be increased more (Figure 1b), but even for 90% reduction of contacts the curve has not been flattened. In Figure 1c the impact of the timing of social distancing measures on the further course of the outbreak is shown. The figure demonstrates how every week delay in implementing these measures leads to an order of magnitude higher numbers of cases, thus emphasizing the importance of fast implementation of social distancing measures. Finally, in Figure 4d we show a scenario where social distancing is implemented at day 28 after diagnosing the first case and reduces non-household contacts by 70%. Then at day 49, also contact tracing is implemented. It is assumed that all infected household contacts and 50% of the non-household contacts of a newly diagnosed case are traced and isolated. This additional measure increases the doubling time to … days. We contrast this scenario with data from epidemics in a number of countries that have implemented social distancing at various levels and time points.

**Figure 1:**
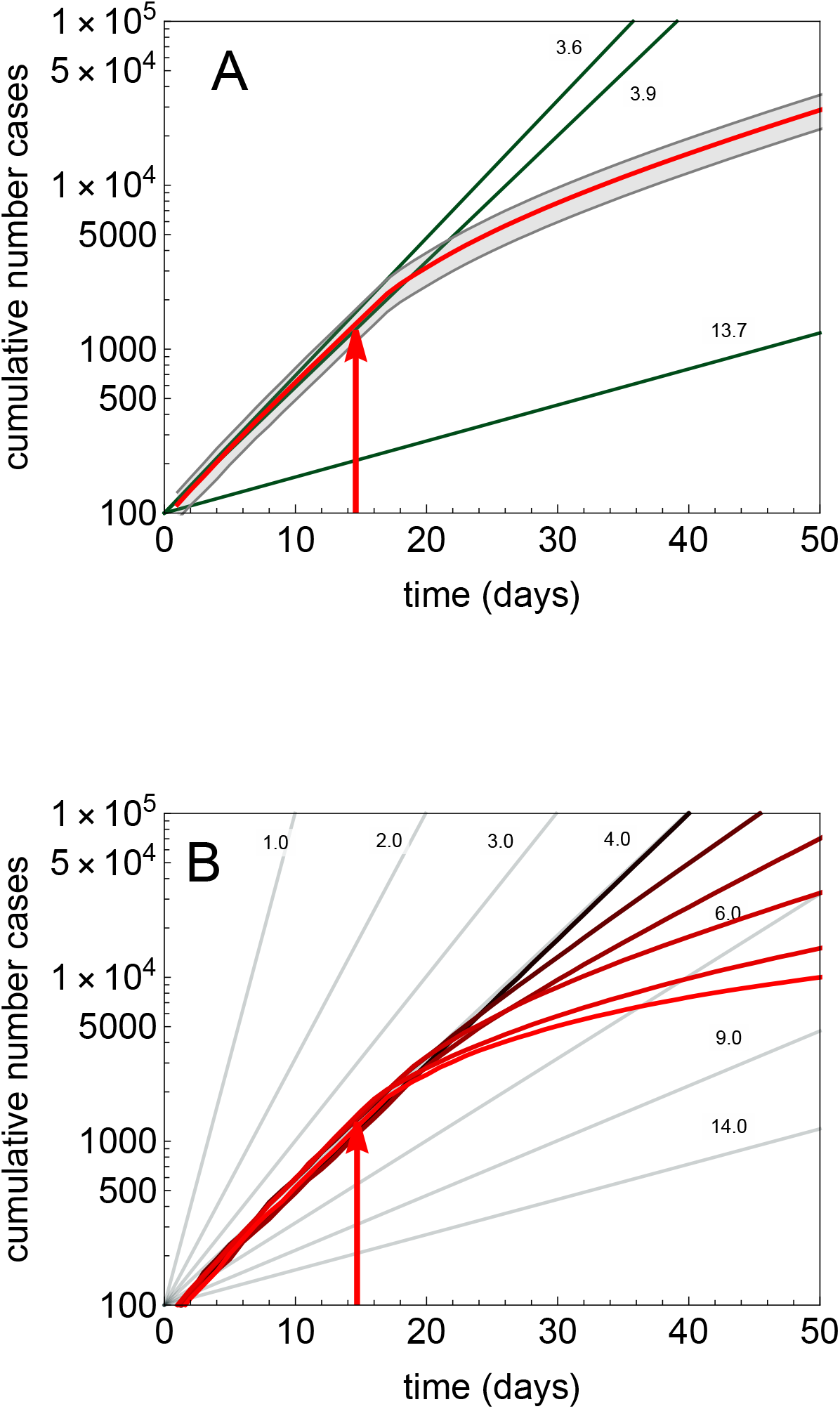
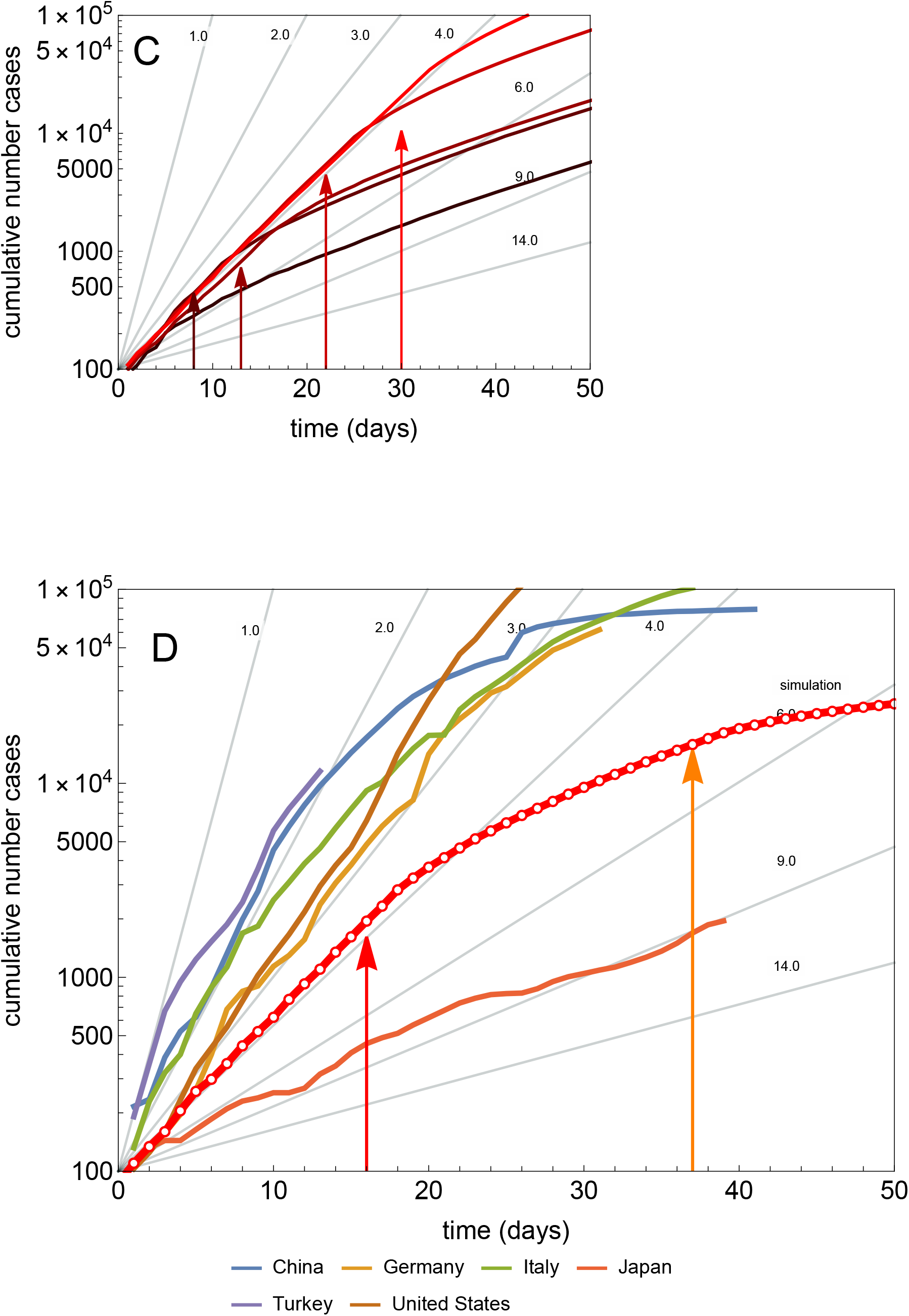
The cumulative number of cases in an outbreak shown from the time when the number of cases exceeds 100. In (A) the mean and standard deviation of 20 simulation runs is shown for a situation where up to day 28 after the first case was diagnosed, only diagnosis and case isolation is implemented. At day 28 after diagnosis of the first case (arrow) social distancing was implemented, which reduced the number of non-household contacts by 70%. The green lines show the doubling times without intervention (3.6 days), with isolation of diagnosed cases (3.9 days), and with social distancing (13.7 days). In (B) simulations are shown with varying levels of social distancing (contact reductions of 0%, 30%, 50%, 70%, 80%, 90%; colors from brown to red; gray lines are doubling times). In (C) social distancing is implemented at the level of 70% reduction at different times after the diagnosis of the first case (2, 3, 4, 5, and 6 weeks, see arrows). In (D) a scenario is shown (red dotted line) with implementation of social distancing at a contact reduction of 70% (red arrow) at day 28; additional contact tracing and isolation and quarantine of contacts starts at day 49 (orange arrow) after diagnosis of the first case. We contrast this scenario with data from outbreaks in several countries that have implemented social distancing at various levels and times during the outbreak.

### Fraction of non-household contacts needed to trace and isolate

If there is a diagnosis delay, the question arises how successful contact tracing has to be to keep the outbreak under control. We therefore compute the minimum fraction of non-household contacts that need to be traced and isolated (henceforth termed “critical tracing coverage”) to bring R_e_ to below 1 (for examples with various assumptions on timeliness of contact tracing see Supplementary Information).

### Impact of asymptomatic cases

Not being diagnosed can be a consequence of not developing symptoms, having only mild symptoms, or any other reason why infected persons might not be identified by health care. We subsume these possible reasons for cases not being ascertained under the term “asymptomatic”. With increasing proportion of asymptomatic cases, the possibility of controlling the outbreak with contact tracing and isolation quickly fades. This is shown in Figure 2, where we plotted the critical tracing coverage for non-household contacts for several values of R_0_ as a function of the fraction of symptomatic cases (i.e. the fraction of those who will eventually develop symptoms during their entire infectious period). Household contacts are assumed to be always traced and isolated. The figure shows that for R_0_ = 2.5 control is not possible with isolation and contact tracing, if less than 60% of all infected persons develop symptoms or are otherwise not detected by the health care system, even if all other parameters are at the most optimistic values. Other interventions measures such as social distancing are then needed for containment of the outbreak.

**Figure 2:**
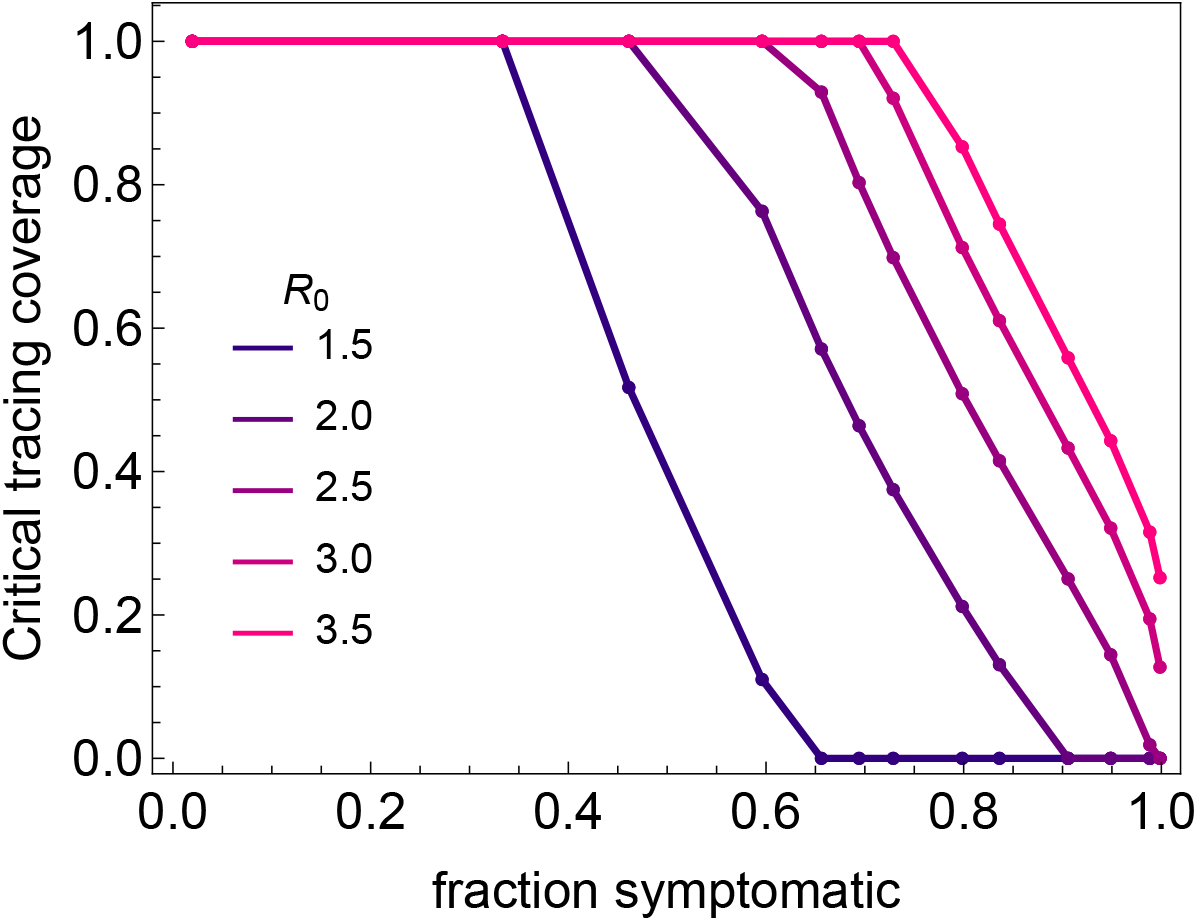
The critical tracing coverage needed for control of the outbreak for varying percentages of asymptomatic infections and values of R_0_ between 1.5 and 3.5. If more than 40% of cases escape diagnosis because they are asymptomatic or have only mild infections, for R0 = 2.5 the outbreak is not controllable even with our optimistic baseline values for the intervention parameters.

### Social distancing and contact tracing

Social distancing in theory can reduce the effective reproduction number to below 1, but only if non-household contacts are almost reduced to zero. In reality this is not possible, because there are many non-household contacts which cannot be avoided to keep essential processes in society going. Therefore social distancing will not be complete and may not be sufficient for containment. Additional tracing and isolation of household contacts may then be the additional effort needed to achieve containment. In figure 3a we show for various levels of social distancing, expressed as proportion reduction of non-household contacts, what fraction of household- or additionally non-household contacts need to be traced and isolated to reduce the effective reproduction number. Similarly, figure 3b shows the increase of the doubling time that can be achieved with these contact tracing measures depending on level of social distancing. Figure 4a shows for various values of R_0_, what are the critical tracing coverages of household contacts only in order to reach an effective reproduction number of 1, which is the threshold to containment. If tracing household contacts is not sufficient, also non-household contacts have to be traced and isolated, for which the critical coverages are shown in figure 4b. For example, if social distancing leads to a reduction of 50% of non-household contacts, additional tracing and isolating household contacts can further reduce the effective reproduction number, but cannot achieve containment. If then also 20% of non-household contacts can be traced and isolated, the transition to containment can be made.

**Figure 3:**
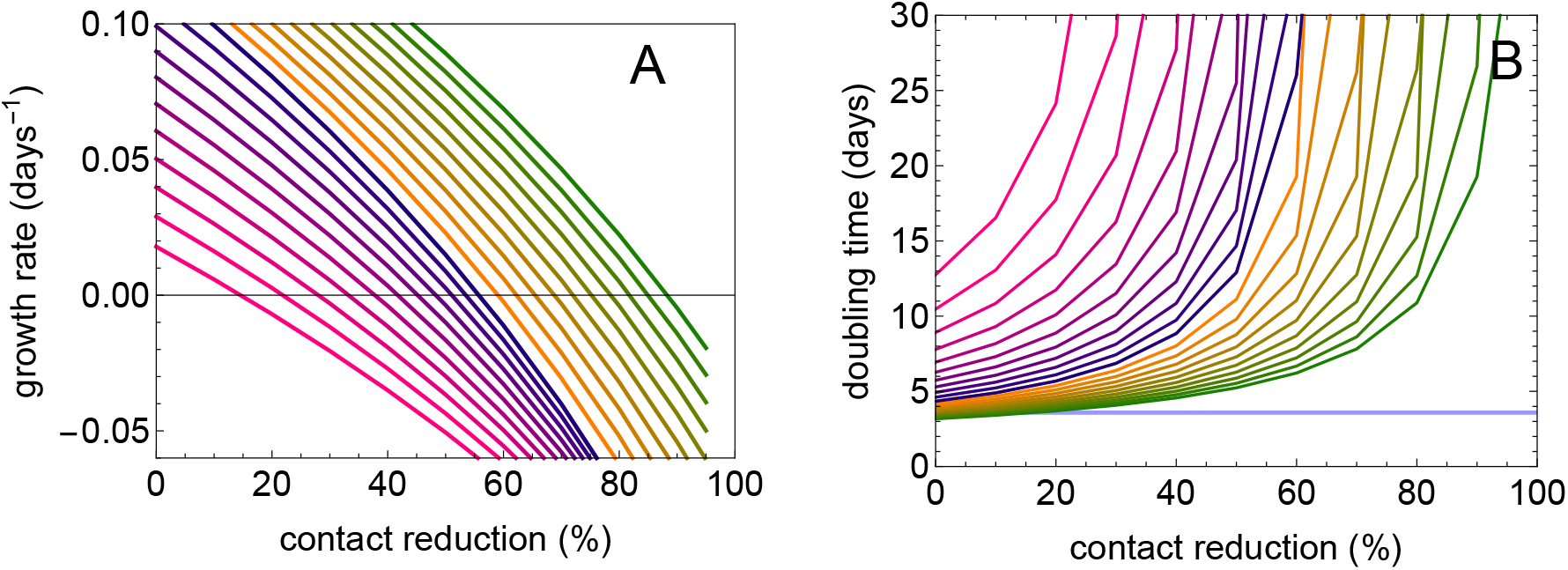
The additional impact of isolation and contact tracing in a population where social distancing is practiced on (A) the exponential growth rate and (B) the doubling time of the epidemic. The additional impact of tracing and isolating household contacts only is shown in the curves from green to yellow (tracing coverage varying from 0% to 100%). If 100% of household contacts are traced and isolated, additional benefit is gained with tracing also non-household contact with increasing tracing coverage shown colored blue to pink (varying from 0% to 100%). In these computations we assumed that 60% of cases are ascertained and R_0_=2.5.

**Figure 4:**
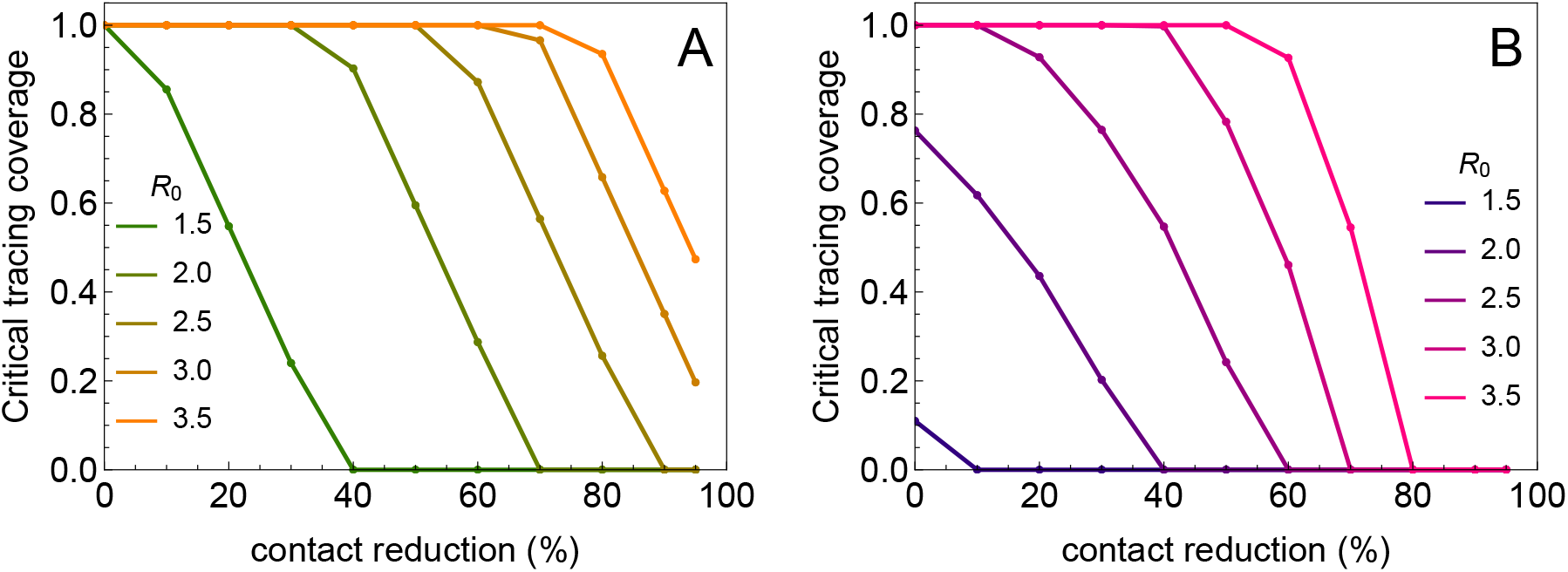
The critical tracing coverages required for containment in a population with social distancing. In (A) the critical tracing coverage is shown if only household contacts are traced and isolated. In (B) the tracing of household contacts is assumed to be 100% and the curves show the additional tracing of non-household contacts needed to achieve containment. Again 60% of cases are assumed to be ascertained and tracing coverages are shown for various values of R_0_.

## Discussion

Our analyses show that rapid diagnosis and isolation of infections based on COVID-19 disease alone cannot control outbreaks of SARS-CoV-2, but that the addition of tracing and isolation of traced cases could in theory be successful depending on the value of R_0_ (Figure 2). In practice, however, the potential for containment will be seriously jeopardized by various delays and imperfections in the tracing process. Especially delays in diagnosis and isolation, and the existence of asymptomatic and mild infections that contribute to onward transmission will make an outbreak uncontrollable. As evidence is mounting that the proportion of asymptomatic and mild cases is large and leads to substantial numbers of unascertained cases [22], most countries have now switched to strategies of social distancing or complete lockdowns. Such measures have proven effective earlier during the 2009 influenza pandemic [23, 24]. However, social distancing can never be complete, as healthcare workers and doctors have to continue their work, but also personnel of supermarkets, public transport employees, and others need to have contact outside their households. Therefore, transmission chains may remain. We find that in a situation where 60% of cases are ascertained, social distancing of non-household contacts fails to contain the epidemic even if contacts outside the household are reduced by 80%. In that case, combining the social distancing with tracing and isolation of household contacts may suffice to bring the balance towards containment. If social distancing is less severe, more intensive contact tracing and also tracing of non-household contacts is needed (Figures 3 and 4). If social distancing reduces non-household contacts only by 50%, tracing and isolation also of non-household contacts is needed for containment. If this is not possible, for example due to constraints of the public health system, tracing and isolation of household contacts can at least substantially increase the doubling time of the epidemic (Figure 3b).

In the recent report by Ferguson and colleagues (https://www.imperial.ac.uk/mrc-global-infectious-disease-analysis/news--wuhan-coronavirus/) it is assumed that non-household contacts would be reduced by 75% by social distancing [25], and a contact study in the UK shows that numbers of contacts are reduced by around 73% [21]. Using these estimates, our model predicts that additional tracing and isolation of infected household contacts would suffice as an additional measure to achieve containment. Possibly, this could be achieved by using digital tracing applications as implemented in South Korea, where it has apparently helped to bring the epidemic under control [3].

Even though the pandemic situation shows that COVID-19 epidemics cannot be contained by contact tracing and rapid isolation alone, this does not render contact tracing useless. On the contrary, contact tracing and isolation when used in addition to social distancing, may be the tool needed to make this mix of strategies successful. Our analyses show that isolation and contact tracing when combined with social distancing can contribute to reducing the growth rate and increasing the doubling time of epidemics, thereby buying time, spreading the number of severe cases out over a longer period of time, and potentially also reducing the total number of infections (the “final size”) [26]. This will lower peak healthcare demand, alleviate the stress on healthcare systems, and contribute to reducing the burden of disease.

Our analyses of contact tracing add to an earlier study by a more systematic analysis of the relation between key parameters (transmissibility, fraction asymptomatic, fraction of contacts traced, diagnosis delays), and by incorporating household versus non-household contacts [27]. Household contacts are at a higher risk of becoming infected than non-household contacts as persons in a household will usually have repeated contacts. On the other hand, our analyses show that household infections contribute less to onward transmission than non-household infections simply because the numbers of household contacts are much lower than numbers of other contacts. As a consequence, the effectiveness in isolating non-household contacts is key for a successful contact tracing strategy.

A strength of the model is that quantitative information about distributions of the latent and infectious periods, and the infectivity per day of the infectious period can be incorporated easily and detailed, such that if new and better data on those quantities emerge, the analysis can be updated quickly. In particular, the model can incorporate non-standard distributions based on empirical data (e.g. viral load measurements to quantify infectiousness per day), should they become available.

A limitation of the analyses presented here is that they apply to a situation in which the epidemic is described by a branching process and is growing exponentially. This also applies to another modelling using a (one-type) branching process.[27] Ultimately, as the number of persons who are or have been infected increases, the number of persons that are still susceptible will start to dwindle, and epidemic growth will ultimately come to a halt. Hence, strictly speaking our results apply to the early stages of an epidemic. In fact, even when the number of infected persons is still relatively small in the early stage of an epidemic it is possible that exponential growth is not observed, for instance due to local depletion of susceptible persons in combination with clustering in contact patterns, spatial effects, and inhomogeneous mixing [28]. However, estimates of the effective reproduction number are independent of the dynamics and give information about the ability of an intervention to stop of slow down epidemic spread. Also, at present the epidemics in many countries are still growing exponentially [19].

In conclusion, our results show that in populations where social distancing is implemented, isolation and contact tracing can play an essential role in gaining control of the COVID-19 epidemic. On their own, none of these strategies are able to contain COVID-19 for realistic parameter settings, but in a combined strategy they can just tip the balance towards containment. These insights provide guidance for policy makers, who will have to decide when and how to release severe lockdown or social distancing measures, and whether additional contact tracing and isolation is then a feasible alternative keep a resurging epidemic at bay.

## Data Availability

We only used data from published literature.

## Contributors

MEK, MvB, and GR conceived the study. MK designed and programmed the model, and produced output. All authors interpreted the results, contributed to writing the manuscript, and approved the final version for submission.

